# COVID-19 Classification of X-ray Images Using Deep Neural Networks

**DOI:** 10.1101/2020.10.01.20204073

**Authors:** Elisha Goldstein, Daphna Keidar, Daniel Yaron, Yair Shachar, Ayelet Blass, Leonid Charbinsky, Israel Aharony, Liza Lifshitz, Dimitri Lumelsky, Ziv Neeman, Matti Mizrachi, Majd Hajouj, Nethanel Eizenbach, Eyal Sela, Chedva Weiss, Philip Levin, Ofer Benjaminov, Gil N Bachar, Shlomit Tamir, Yael Rapson, Dror Suhami, amiel a dror, Naama Bogot, Ahuva Grubstein, Nogah Shabsin, Yishai M Elyada, Yonina Eldar

## Abstract

**Objectives:** In the midst of the coronavirus disease 2019 (COVID-19) outbreak, chest X-ray (CXR) imaging is playing an important role in diagnosis and monitoring of patients with COVID-19. Machine learning solutions have been shown to be useful for X-ray analysis and classification in a range of medical contexts. In this study, we propose a machine learning model for detection of patients tested positive for COVID-19 from CXRs that were collected from inpatients hospitalized in four different hospitals. We additionally present a tool for retrieving similar patients according to the model’s results on their CXRs.

**Methods:** In this retrospective study, 1384 frontal CXRs, of COVID-19 confirmed patients imaged between March-August 2020, and 1024 matching CXRs of non-COVID patients imaged before the pandemic, were collected and used to build a deep learning classifier for detecting patients positive for COVID-19. The classifier consists of an ensemble of pre-trained deep neural networks (DNNS), specifically, ReNet34, ReNet50, ReNet152, vgg16, and is enhanced by data augmentation and lung segmentation. We further implemented a nearest-neighbors algorithm that uses DNN-based image embeddings to retrieve the images most similar to a given image.

**Results:** Our model achieved accuracy of 90.3%, (95%CI: 86.3%-93.7%) specificity of 90% (95%CI: 84.3%-94%), and sensitivity of 90.5% (95%CI: 85%-94%) on a test dataset comprising 15% (350/2326) of the original images. The AUC of the ROC curve is 0.96 (95%CI: 0.93-0.97).

**Conclusion:** We provide deep learning models, trained and evaluated on CXRs that can assist medical efforts and reduce medical staff workload in handling COVID-19.

**Key Points:**

- A machine learning model was able to detect chest X-ray (CXR) images of patients tested positive for COVID-19 with accuracy and detection rate above 90%.
- A tool was created for finding existing CXR images with imaging characteristics most similar to a given CXR, according to the model’s image embeddings.

## 1 Introduction

The Coronavirus Disease 2019 (COVID-19) pandemic, caused by the SARS-CoV-2 virus, poses tremendous challenges to healthcare systems around the world, and requires physicians to make fast clinical decisions under pressure. After many months that led to exhaustion of the medical teams, hospitals are confronting renewed surges with overwhelming numbers of new patients seeking medical aid. Some patients approach the emergency departments with respiratory symptoms, and others that are being evaluated for different reasons, are asymptomatic yet positive for COVID-19.

The prevalent test used for COVID-19 identification is Reverse Transcription Polymerase Chain Reaction (RT-PCR) (1–3), despite its high false negative rates. The undetected fraction of active patients inevitably leads to uncontrolled viral dissemination, masking hidden essential epidemiological data (4–6). Additionally, RT-PCR testing kits are expensive, processing them requires dedicated personnel and can take hours to days. Rapid and accurate methods of diagnosis that do not rely on medical staff are therefore becoming crucial for the control of the pandemic. CXRs of COVID-19 patients can demonstrate typical findings including peripheral opacities and ground glass patterns in the absence of pleural effusion (5,7,8), and therefore may be used as a triage test, for establishing and grading pulmonary manifestations, as well as for follow up.

Deep learning models have shown impressive abilities in image related tasks, including in many radiological contexts (9–11). They have great potential in assisting COVID-19 management efforts, but require large amounts of training data. When training neural networks for image classification, images from different classes should only differ in the task specific characteristics; it is important, therefore, that all images are taken from the same machines. Otherwise, the network could learn the differences, e.g., between machines associated with different classes rather than identifying physiological and anatomical COVID-19 characteristics.

Portable X-ray machines are predominant in COVID-19 handling (12), and most available CXRs of patients with COVID-19 in Israel come from portable X-rays. While COVID-19 is easier to detect in CT (13), CT is more expensive, exposes the patient to higher radiation, and its decontamination process is lengthy and causes severe delays between patients. The major challenge with the use of CXR in COVID-19 diagnosis is its low sensitivity and specificity in current radiological practice. A recent study found that the sensitivity of CXRs was poor for COVID-19 diagnosis (14).

This study aims to develop and evaluate machine learning tools for COVID-19 identification and management. A large dataset of images from portable X-rays collected in 4 different hospitals was used to train and evaluate a network that can detect COVID-19 in the images with high reliability and to develop a tool for retrieving CXR images that are similar to a query CXR image, based on a metric defined by the classifier. The network results in detection accuracy of 90.3%, specificity of 90% and sensitivity of 90.5%.

## 2 Materials and Methods

### Data and patients

This retrospective study took place during and after the first wave of the COVID-19 pandemic in Israel, and included patients aged 18 years and older in four medical centers in Israel. The data for this study includes a total of 2427 frontal (AP/PA) CXR images from 1384 patients (63 +/-18 years, f:m= 832:552), 360 of which with a positive COVID-19 diagnosis and 1024 negative. All images come from portable X-ray machines. For COVID-19 positive patients, the standard protocol was that every symptomatic patient with positive RT PCR test for COVID 19 was admitted to the hospital, even if symptoms were mild. Routine chest X-rays were performed at the day of admission and then later for follow up. COVID-19 positive images include a wide range of minimal to severe pulmonary damage, which, for the purpose of this work were all read as positive COVID-19. The non COVID-19 images were obtained from CXRs taken by the same X-ray machines from January 2017 to April 2019, before the start of the pandemic, meaning there are no false negatives in our cohort. These include normal as well as abnormal radiographs with other clinical conditions.

The test set was taken from the full CXR dataset and contains 350 CXR (15%) of which 179 (51%) are positive for COVID-19 and 171 (49%) are negative. For patients with multiple images, their images were used either for the test set or for the train set, never for both. This is done to prevent the model from identifying patient-specific image features (e.g., medical implants) and associating them with the label. Both train and test sets include patients from all four hospitals.

All images were used in the highest available resolution without lossy compression; 4% (101/2426) of the images were excluded due to lateral positioning, or due to rectangular artifacts in the image, of these 98 were COVID-19 positive. No additional selection criteria were used to exclude images based on clinical radiological findings.

### Image Processing

The model pipeline (Figure 1), begins with a series of preprocessing steps, including augmentation, normalization, and segmentation of the images.

**Figure 1:**
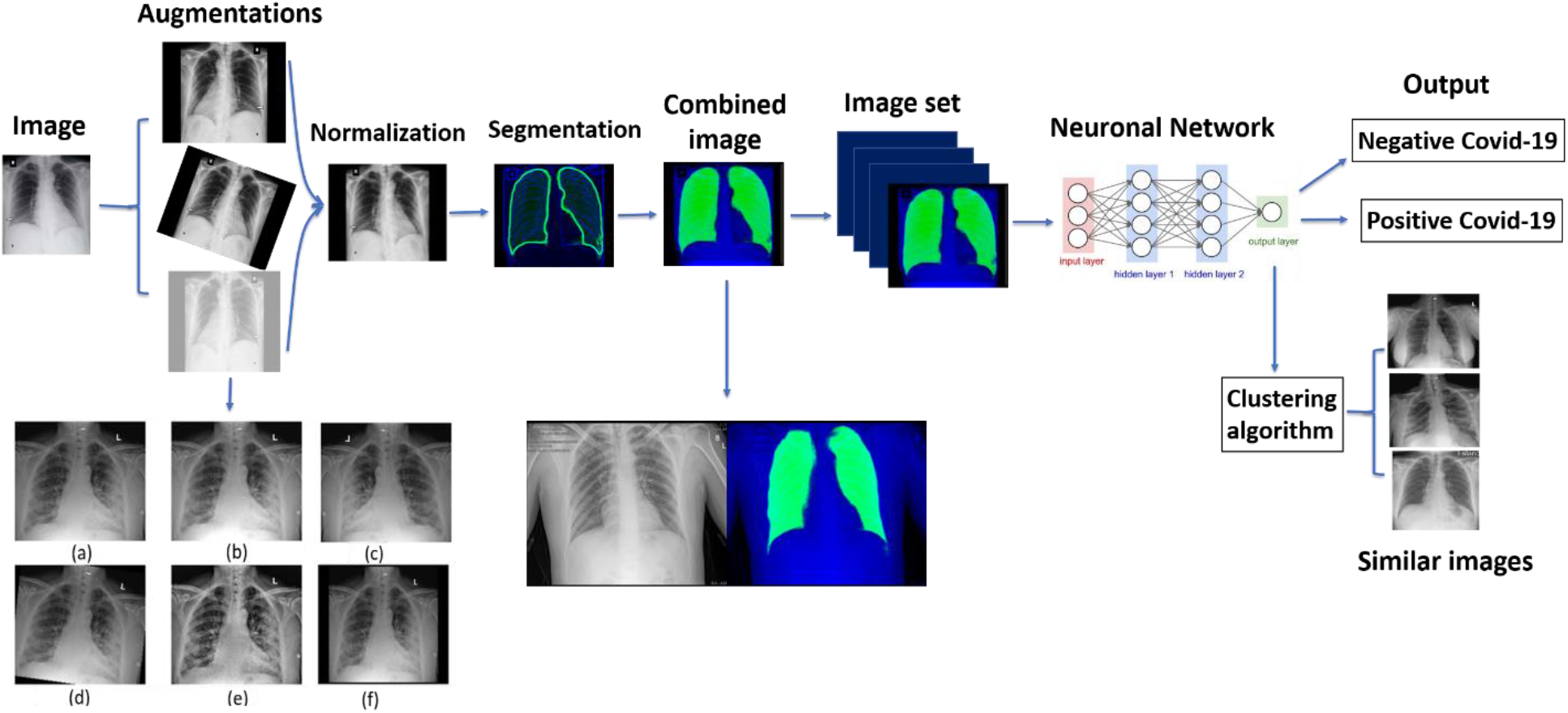
Full pipeline workflow overview. First each image undergoes processing consisting of: augmentation, which is a set of visual transformations (transformations shown: (a) original image, (b) brighten, (c) horizontal flip, (d) 7 degrees rotation, (e) CLAHE transformation, (f) scale), normalization, in order to set a standard scale of image size and color, and segmentation, which emphasizes the area of the lungs and is combined to the image. The entire image set is then fed into a Neural Network which produces a classification outcome for each image as positive for coronavirus disease 2019 (COVID-19) or negative for COVID-19. In addition, embedded features are extracted from the last layer of the network and are used to find images with similar characteristics to a given image as learned by the network.

Augmentations are transformations that change features such as image orientation and brightness. These properties are irrelevant for correct classification, but may vary during image acquisition, and can affect the training performance of the network because of its rigid registration with respect to orientation and pixel values. They serve to enlarge the dataset by creating a diverse set of images, increasing model robustness and generalizability (15,16). Importantly, augmentations should correspond to normal variation in CXR acquisition; to ensure this we consulted with radiologists when defining the augmentation parameters (see Supplemental Material for details).

The normalization process aims to standardize image properties and scale. It consists of cropping black edges, standardizing the brightness and scaling the size of each image to 1024×1024 pixels using bilinear interpolation.

To enhance performance we created an additional image channel using lung segmentation via a U-net pre-trained on an external dataset as detailed in (17). This network produces a pixel-mask of the CXR indicating the probability that each pixel belongs in the lungs, allowing the network to access this information while training. Input images contain 3 channels: the original CXR, the segmentation map, and one filled with zeroes. This is done to accommodate the pre-trained models we used that use 3-channel RGB images.

### Network architecture and output

We compared five network models: ResNet34, ResNet50, ResNet152 (18), VGG16 (19) and CheXpert (9). The general approach of these architectures is to reduce images from a high-dimensional to a low-dimensional space such that a simple boundary can be used to separate image classes. The models were trained using transfer learning, i.e. loading weights pre-trained on the ImageNet database (18,19) or on the CheXpert dataset (9) and subsequently retraining them on our data. We additionally classify the images using an ensemble model that outputs the average of the networks’ results.

In addition to classification, we propose a method for retrieving a number of CXR images that are the most similar to a given image. The activation of layers of the neural network serve as embeddings of the images into a vector space, and should capture information about clinical indications observed in the images. We use the embeddings produced by the network’s last layer to search for similarity between the resulting vectors, and retrieve the nearest neighbors of each image.

### Statistical analysis

For model evaluation we used accuracy, sensitivity, specificity, and area under the curve (AUC) for receiver operating characteristic (ROC) and precision recall (P-R) curves. Confidence intervals (CIs) were calculated by running a training of the model 10 times using different divisions of training and testing sets, then taking 100 bootstrap samples out of the test set for each division, and calculating the requested metrics on each of these divisions and bootstrap samples. The CIs for each run are then given by the 2.5th and 97.5th percentiles for each metric. We report the CIs for the original data split within the paper. See Supp. Fig s7 for more detailed results from all 10 data divisions.

## 3 Results

### Data acquisition

The patient data included in this study are shown in Table 1. The imaging dataset consists of a total of 2426 CXRs, of which 53% (1289/2426) are positive for COVID-19 and 47% (1138/2426) are negative; 4% (101 of 2426, 98 positive) of the images were excluded due to lateral positioning or having rectangular artifacts covering parts of the image. To our knowledge this is one of the largest datasets of original COVID-19 labeled X-ray images.

**Table 1:**
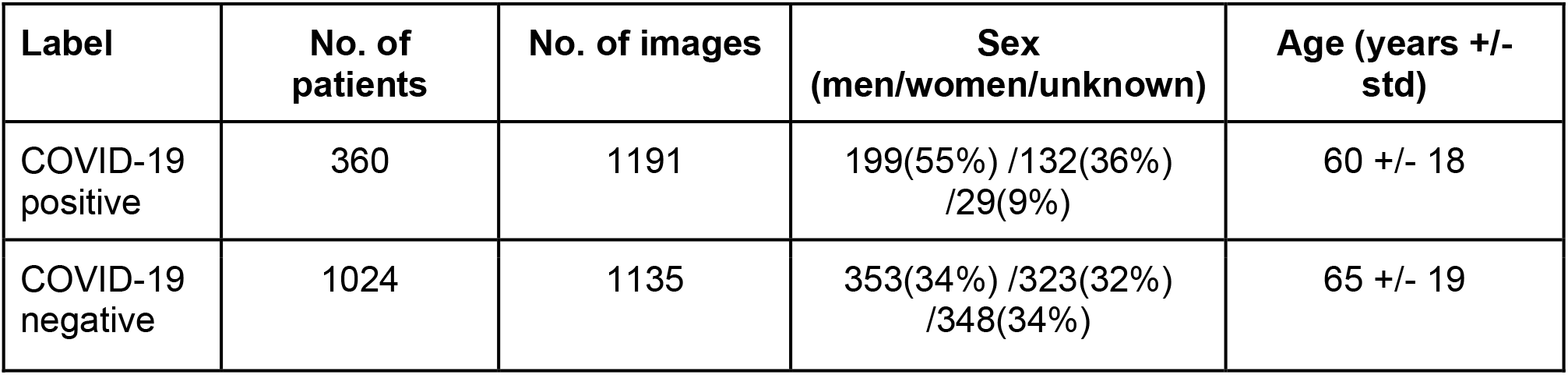
**Demographic statistics on patients and chest images in this study**.

### Quantitative analysis of the model

The performance of the network was tested on 15% (350 of 2426) of the images that were not used for training. The metrics we used are accuracy (proportion of successful classifications), sensitivity (also – recall, the proportion of positively labeled images that were classified correctly) and specificity (proportion of correctly classified negative images). Results for five different networks can be seen in Table 2. The neural network with the best results was ResNet50, and was used for analyses requiring network embeddings (t-SNE and KNN). The ensemble model, which averages over the output of multiple networks, achieved accuracy of 90.3%, (95%CI: 86.3%-93.7%) specificity 90.0% (95%CI: 84.3%-94%), and sensitivity 90.5% (95%CI: 85%-94%) on the test images. The AUC of the ROC curve is 0.96 (95%CI: 0.93-0.97). The ROC curve is provided in Figure 2a, showing the relationship between the false positive rate (FPR) and the true positive rate (TPR) for different classification threshold values. Figure 2b presents the P-R curve, which shows a similar tradeoff between precision (proportion of positively classified images that were correctly classified) and recall, with AUC of 0.96 (95%CI: 0.94-0.97). Both figures show a broad range of thresholds for which both high performance metrics are attainable.

**Table 2:**
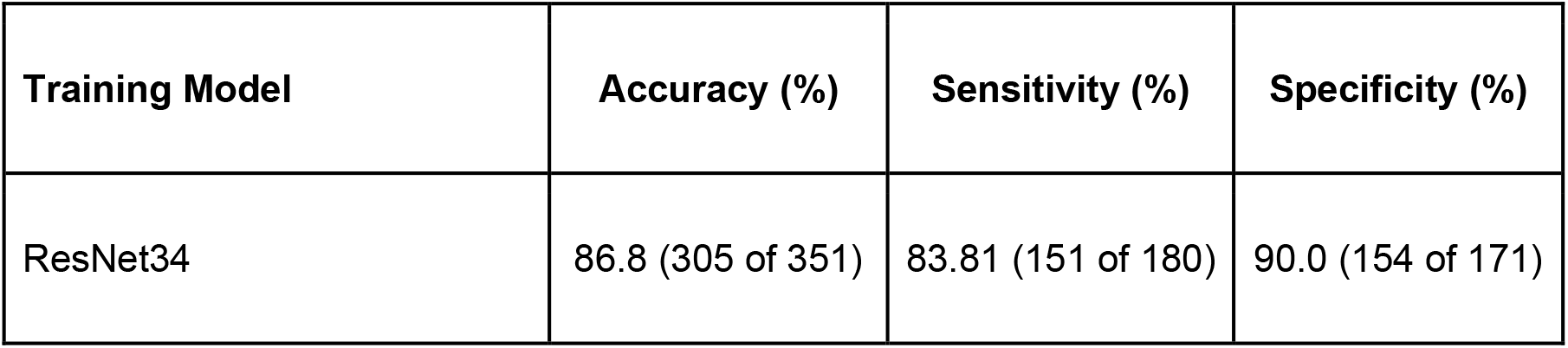
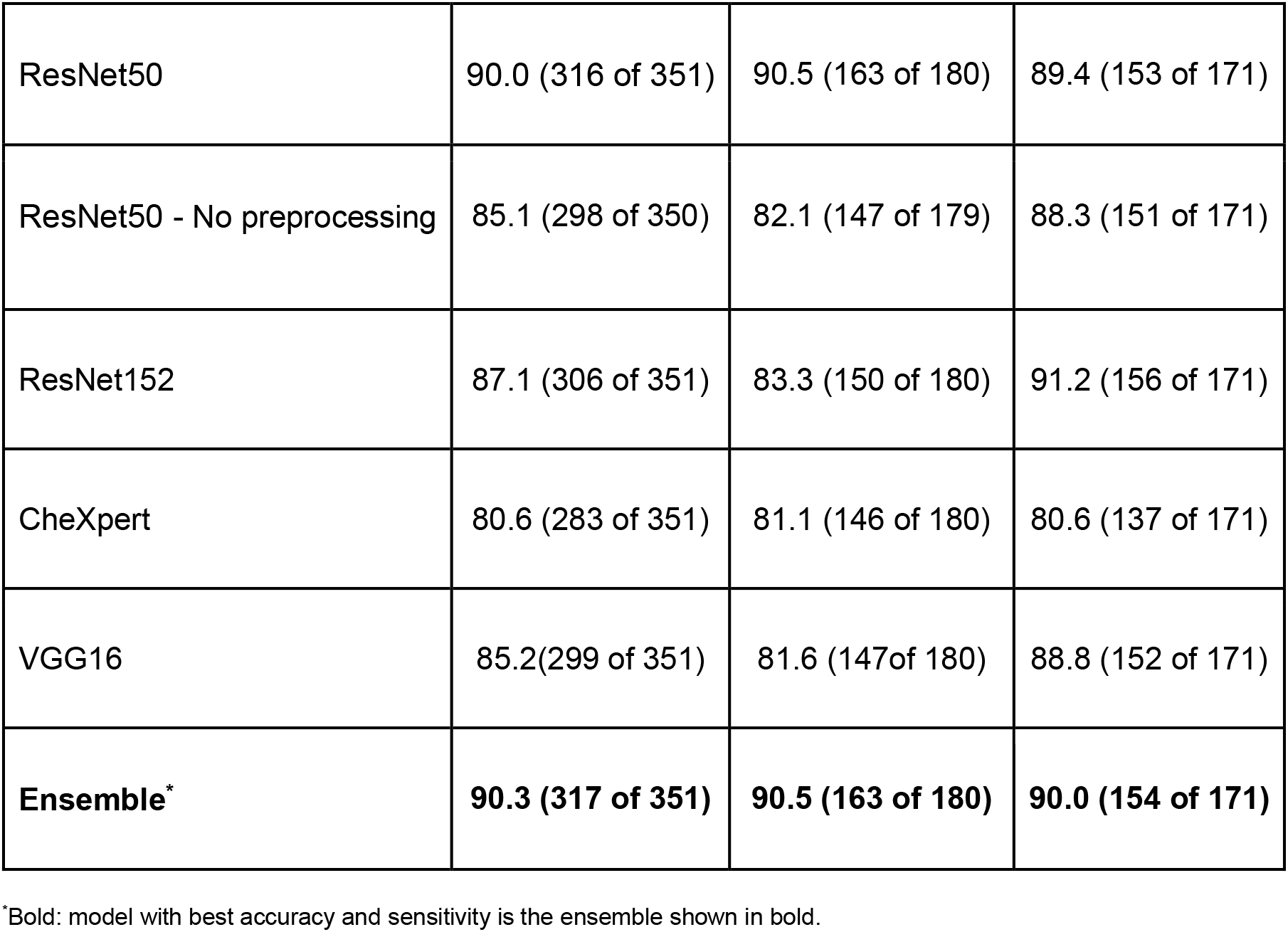
**Comparison of accuracy, sensitivity and specificity of various deep networks trained and tested on the same test set**.

**Figure 2:**
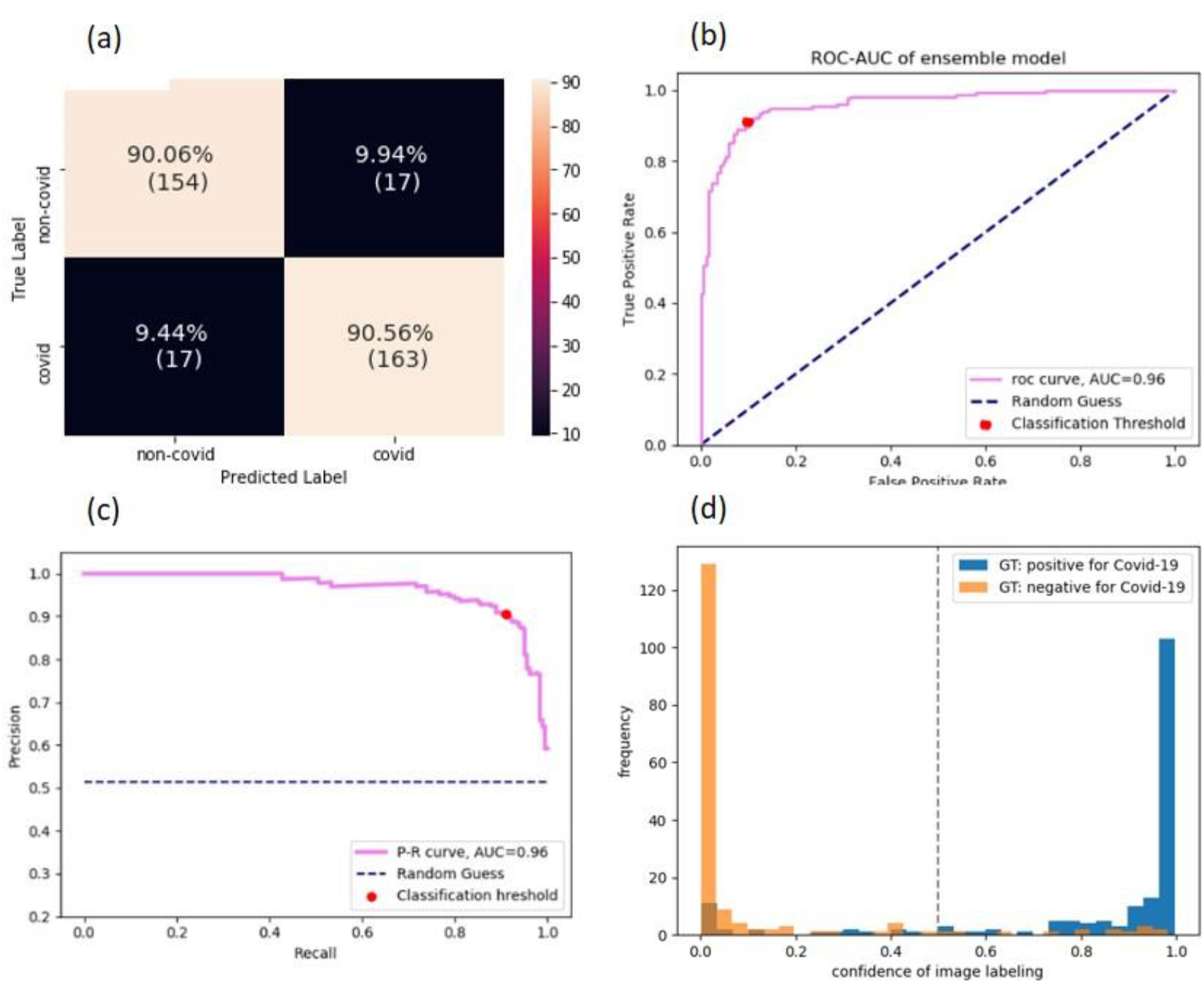
Performance of the model. (a) Confusion Matrix of the classification. True positive rate (TPR) at the bottom right corner, true negative rate (TNR) at the top left corner, false positive rate (FPR) at the top right corner, and false negative rate (FNR) at the bottom left corner. (b) Receiver Operating Characteristic (ROC) curve. The curve shows the relation between true positive rate (TPR) and false positive rate (FPR) as the threshold of the separation between positive and negative classification is varied. The performance of the model is measured by the area under the curve (AUC). Ideally, the curve should cover as much area as possible up to the upper left corner (AUC score of 1), which minimizes the FPR while maximizing the TPR. The AUC is 0.95 (c) Precision-Recall curve. Shows the relation between Precision and Recall. Precision and Recall are affected from different classes of the data, thus can vary in scores when data is imbalanced (e.g. more observations of positive or negative compared to the other). We would like to have the AUC as large as possible up to the upper right corner, which maximizes both Precision and Recall. (d) Classification score histogram. Ground truth (GT) labels are in colors. Every image is scored on a scale between 0 and 1 with threshold of 0.5, seen as a dashed line, such that all images with a higher score will be classified as positive for COVID-19 and images below as negative. Negatively labeled images that received a score above 0.5 are, therefore, incorrectly classified images, and vice versa with respect to positively labeled images. However, the closer the image score is to one of the edges (0 or 1), the stronger the confidence in the image’s classification. The accumulation of two distinct colors on the edges point to good separation of many observations with strong confidence in the classification.

We trained the ResNet50 model on the dataset with and without all preprocessing stages. As seen in Table 2, preprocessing incurs an improvement of 4% in accuracy and 5% in sensitivity. In analyzing subgroups of our patient cohort, we found that prediction accuracies are higher for females than males (Supp. Fig. 1), but there is no strong effect of age on model performance (Supp. Fig. 2).

### Qualitative analysis of the model

The binary decision of whether a patient has COVID-19 is based on an activation score between 0 and 1 outputted by the network and corresponding to the probability the network assigns to the positive label. We generated a histogram of these scores (Figure 2d), and observe that the majority of the correctly classified points are accumulated at the edges, while the wrongly classified images are more spread out along the x-axis.

We additionally visualize the distinction made by the model using t-distributed Stochastic Neighbor Embedding (t-SNE) (20). t-SNE uses a nonlinear method to reduce high dimensional vectors into two dimensions, making it possible to visualize the data points and reveal similarities and dissimilarities between them. We the last layer of the network to obtain an embedding of the images into a vector space. These embeddings are then inputted to the t-SNE. Figure 3 shows these image embeddings as points in a 2-dimensional space, colored by their GT labels. The figure depicts two distinct clusters, revealing a similarity between most images belonging to the same label.

**Figure 3:**
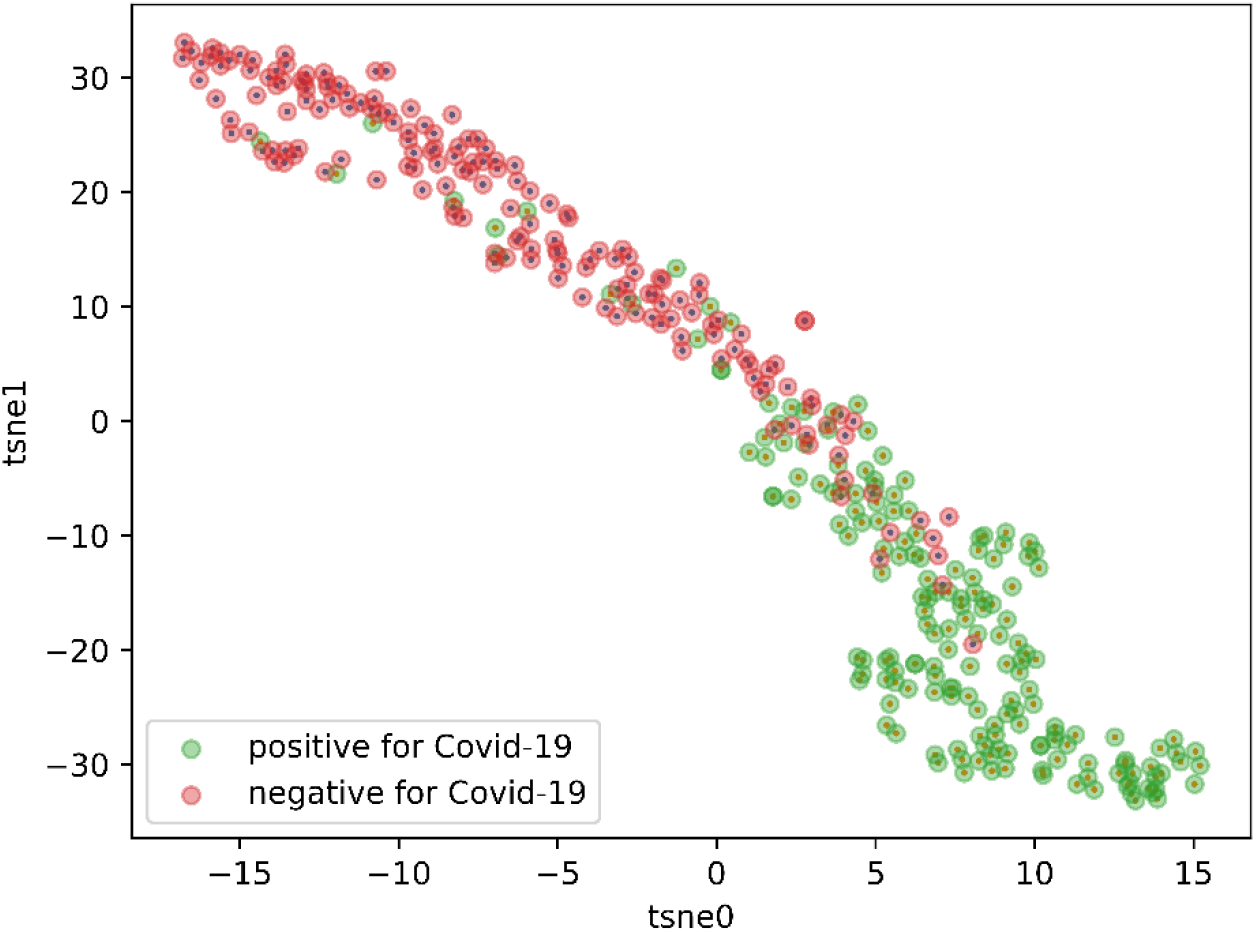
t-distributed Stochastic Neighbor Embedding (t-SNE). A high-dimensional feature vector is extracted for each image from the last layer before the network output, which are used for decision of the output of the neural network, and is reduced into 2 dimensions. Each point on the graph represents the features of an image after dimension reduction and arrangement in space. Next the images were colored according to their ground truth (GT), thus revealing two main clusters. The clusters are mostly in one color each, which essentially shows a strong association of the features, extracted from the decision layer and are used to arrange in space, with the GT of the images, represented by the colors.

We also examine the model’s performance over time, by plotting the prediction scores according to the days from admission. As the disease progresses, lung findings can become more prominent. This is in line with the results, seen in Figure 4. The model performance improves over time, with most classification errors occurring on a patient’s first image, taken upon hospital admission.

**Figure 4:**
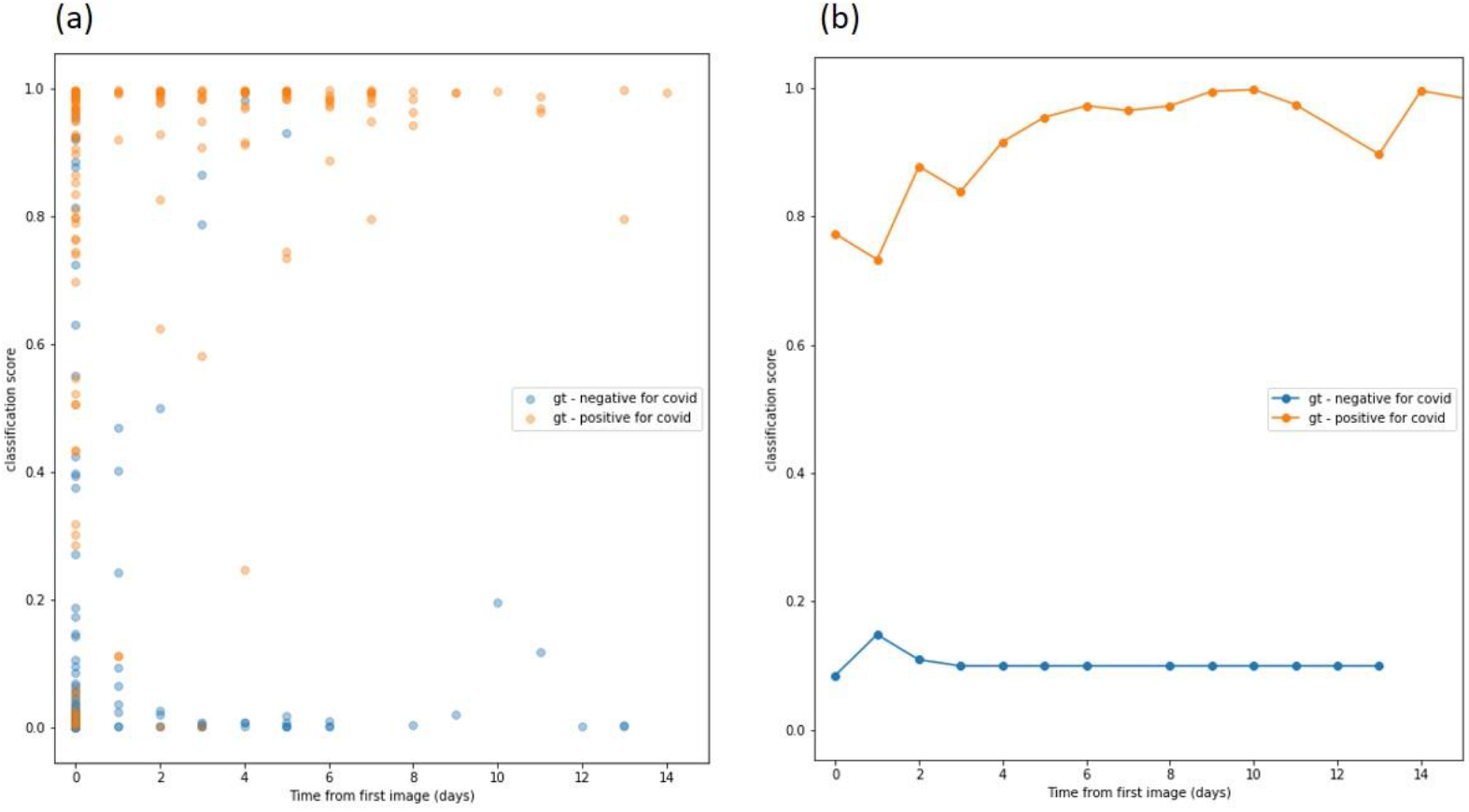
Classification score as a function of time change. The first image of each patient was acquired at the same day of first admission; we note that time value as day 0. Other images of patients which were scanned more than once were noted with time value according to the number of days since the first image was acquired, thus representing the time elapsed from first admission and is ordered on the x-axis. The y-axis shows the classification score of each image between 0 (=negative for COVID-19) and 1(=positive for COVID-19), such that closer to the edge indicates more confidence in the network’s classification. (a) the arrangement of the classification score as a change of time. As more days elapse since first admission, the more confident the classification. (b) Mean values of classification scores for all images of the same day value.

In order to test the model’s performance on a more difficult task, we use it to classify 22 CXRs, 9 positive for COVID-19 and 13 negative, determined by radiologists as challenging to diagnose. Challenging images included images from patients with a positive COVID-19 PCR that either have minimal parenchymal abnormalities and look normal to the radiologist’s eye, or have pulmonary infiltrates similar to preexisting diseases other than COVID 19. The accuracy on this task was 77%, and the sensitivity 77%. In Figure 5, three correctly classified images from this test are shown with the network’s classification score and the GT.

**Figure 5:**
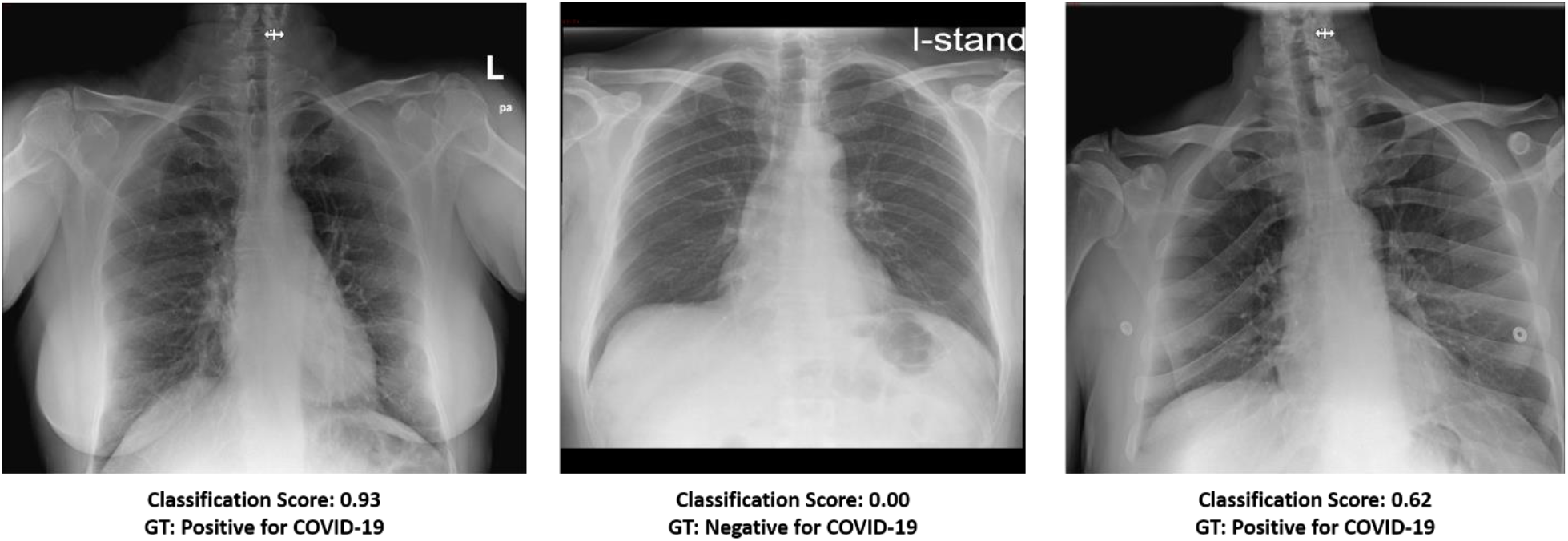
Three images labeled by a radiologist as hard to diagnose. Despite this, the model was able to classify them correctly. Each image is scored with classification score on a scale between 0 and 1 with threshold of 0.5 such that all images with confidence score above the threshold will be labeled as positive for COVID-19 and images below as negative. The ground truth (GT) of each image is also shown.

As an additional tool, we applied K-Nearest Neighbors (KNN) on the image embeddings in order to retrieve images similar to each other as shown in Figure 6. For each image we retrieve 4 images with the closest image embeddings; averaging over these images’ predictions achieves 87% accuracy (305/350), 91.2% specificity (156/171), and 83.2% sensitivity (149/179), meaning that the nearest images typically have the same labels.

**Figure 6:**
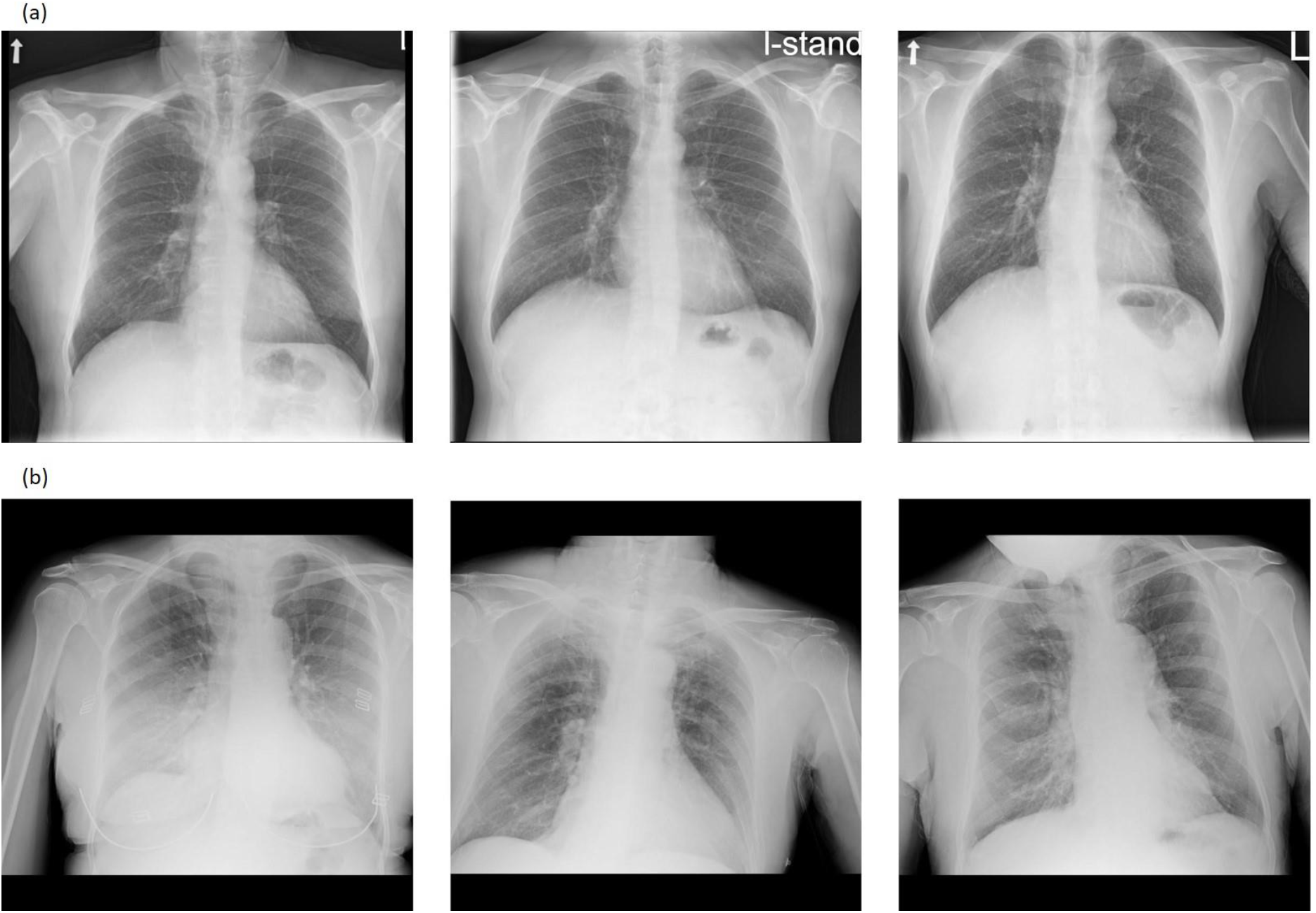
In the figures, the left image is a CXR from the test set, and the two on the right are the two images closest to it from the training set, given the image embeddings from the network’s last layer. (a) All three images are COVID-19 Negative. The distances between the middle and rightmost images to the left one are 0.54 and 0.56 respectively. (b) All three images are COVID-19 positive. The distances between the middle and rightmost images to the left one are 0.51 and 0.55 respectively. The overall mean distance between training and test images is 3.9+/-2.5 (mean +/-std). The mean distance between all positive training and positive test images is 1.4+/-1.9, between negative training and negative test images 2.2+/-1.3, and between images from different classes is 5.8 +/-1.9. Images from different classes are further away from each other, but whether a close distance truly corresponds to similar lung findings still requires verification.

## 4 Discussion

In this study we developed a deep neural network pipeline to classify chest X-ray (CXR) images of patients as coronavirus disease 2019 (COVID-19) positive or negative, and to identify which X-ray scans are similar to each other. We achieved a detection rate of above 90% using both the ensemble model and ResNet50. In addition, we created a tool that retrieves the CXR images most similar to a given image. This can provide physicians with a reference to previous patients that had similar CXR findings. They can use the internal information they have from the hospital about these previous patients to support decisions for further treatment.

Early approaches to COVID-19 classification using neural networks relied on publicly available image sources, including COVID-19 image data collection (21) with 481 COVID-19 positive images and COVID-Net open source initiative with 473 COVID-19 X-ray images (22–26). Some efforts include separation of multiple classes of lung and chest conditions including COVID-19 (27), and others attempt outcome prediction (28,29).

Such efforts have a number of drawbacks, highlighted in the detailed review presented in (30). These datasets were compiled from various sources, often using one source only for COVID-19 images and another only for COVID-19 negative images and other non-COVID conditions (30). Positive and negative images in these datasets may therefore be produced by different X-ray machines, in particular portable and fixed machines, which give rise to images with different expressions of acquisition-related features. This can allow the network’s predictions to rely on features related to the source more than on the relevant medical information (32). In addition, they include a limited number of positive COVID-19 CXR images, which may cause the model to overfit (31), as it is exposed to a relatively small number of characteristics from the data. This impairs the ability to generalize to external datasets. These models’ reliability still needs to be verified on external data. A dataset with more positive COVID-19 images as used in this study, containing 1191 positive CXRs, tends to be more stable.

In this work we sought to address the limitations of previous studies in several ways. Importantly, we took care to include in our dataset CXRs from the same machines both for patients positive and negative to COVID-19. We used raw images without compression that may result in loss of features and introduction of source-dependent artifacts. Moreover, our dataset contained diverse data from four medical centers and was balanced between COVID-19 and non-COVID-19 images.

A recent effort has shown more reliable results based on a larger, more uniformly sourced, dataset and comes closer to the goal of developing tools that can be used in clinical settings (11). They achieved a sensitivity of 88% with a specificity of 79%. Our approach improves on these notably solid results in terms of performance (sensitivity of 90.5% and specificity of 90.0%). As we show, this performance increase may have resulted from the image pre-processing, particularly the inclusion of augmentations and the addition of a segmentation channel. This leads to a performance increase of 8.4 percentage points in sensitivity and 1.1 percentage points in specificity (Table 2 – ResNet50 vs. ResNet50 no preprocessing), and also in balancing of the sensitivity and specificity results.

Another novelty of our work is that we introduced a content-based image retrieval tool that identifies similar CXRs based on a metric defined by using the image embeddings given by the second to last layer of ResNet50. As ResNet50 was trained for COVID-19 classification, we expect similar images under this metric to represent similar cases in terms of their clinical condition. This tool enables medical staff to search the database to identify relevant study cases for a new case under consideration. We note that the scoring process for this similarity measure still requires further investigation in a clinical setting. We would ideally like to compare the disease progression for patients that were found by our tool to have similar lung findings.

As future work, we intend to deploy our model for testing in a clinical setting within the hospitals. We also plan to work on COVID-19 severity classification. A limitation of our study is that preexisting medical illnesses and comorbidities were not integrated into the analysis of both COVID-19 and control datasets, due to a lack of access to clinical data of the patients. Our COVID-19 negative cohort comprises patients with a multitude of diseases, but with the absence of precise labels, we cannot analyze our ability to separate between COVID-19 and any specific lung morbidity. Moreover, our classifier is tailored towards portable X-rays within the four hospitals that provided the data. It requires further fine tuning to be used in other hospitals or diagnostic settings.

In summary, we developed a deep neural network which is able to reliably detect patients with coronavirus disease 2019. Even though medical imaging has not yet been approved as a standalone diagnosis tool (12), we believe it can be used as an aid to medical judgement with the advantage of immediate outcome. We also created a tool for X-ray image retrieval based on lung similarities. This tool can help physicians draw connections between patients with similar disease manifestations, by referring them to images with similar lung characteristics. These images may be linked internally to the corresponding patients, and the treatment and outcome of these patients can then inform decisions upon treatment for the current patient.

## Data Availability

The code will be made available upon publication of the paper

https://github.com/X-ray-Covid-19-AI/covid19_xray_pub

## Abbreviations

CXR: chest x-ray
COVID-19: Coronavirus Disease 2019
RT-PCR: reverse transcription polymerase chain reaction
ROC: receiver operating characteristic
P-R curve: precision-recall curve
AUC: area under the curve
GT: ground truth
FPR: false positive rate
TPR: true positive rate

## Supplemental Material

The supplemental material includes a number of additional analyses. We provide extra details about the performance of the algorithm when considering the sex and age of the subjects, more information about the processing of images and the details of the augmentation procedures, and an additional figure demonstrating the segmentation algorithm. We also provide more details about the architectures used in our study and choice of training parameters. Finally, we provide a bootstrap analysis in order to estimate the dependency of the performance of the model on different divisions of the data.

### Analysis of patient subgroups

The figures below depict the gender distribution and age distribution of the patients in the test set.

**Figure s1:**
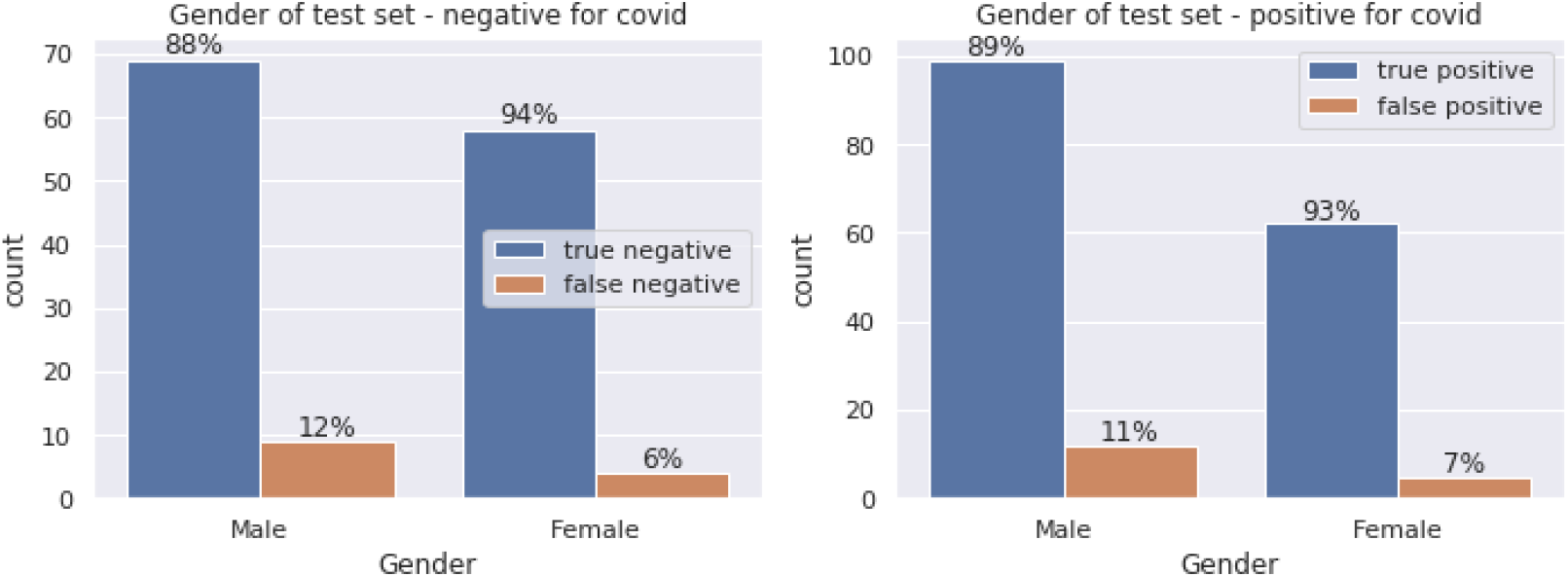
gender distribution of patients in test set (a) Gender of the patients negative for COVID-19. (b) Gender of the patients positive for COVID-19.

**Figure s2:**
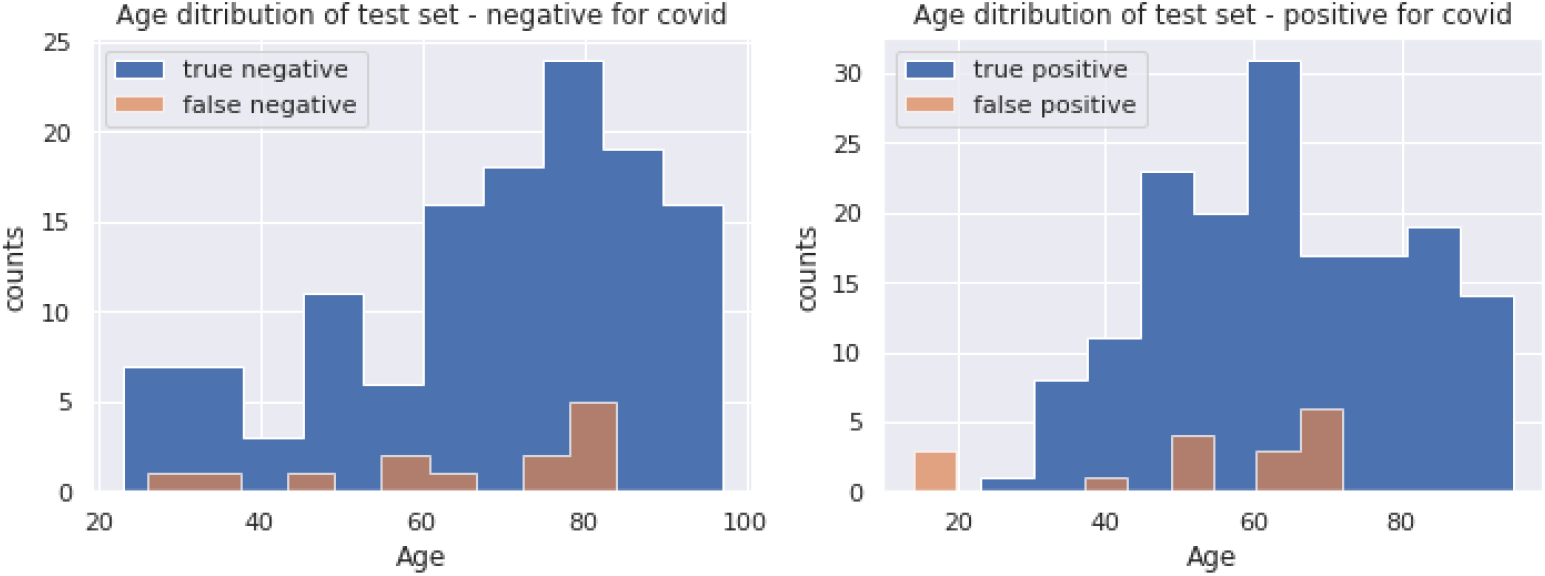
Age distribution of patients in test set. (a) Age distribution of non COVID-19 patients, split by correct and incorrect labels (b) Age distribution of the patients positive for COVID-19.

For two of the hospitals, we have labels specifying whether the lung scan is normal or abnormal for non-COVID-19 patients, abnormal meaning there is some medical finding in the lungs.

| Hospital | Total COVID-19 negative images | Normal | Abnormal |
| --- | --- | --- | --- |
| Rabin Medical Center | 767 | 366 | 401 |
| Shaare Zedek | 145 | 70 | 75 |

### Data preprocessing

Before training, each image goes through a preprocessing pipeline. We start by cropping out areas that contain only text around the images themselves. We then unify the image sizes, preserving the original aspect ratios via padding, and apply a CLAHE filter (a filter that was seen to enhance images and improve deep learning performance (34)). On the training data, we also apply a series of augmentations.

### Augmentation

Augmentations are transformations performed on the data that serve a dual purpose. First, applying the augmentations creates additional diverse sets of images from the existing ones and enables one to artificially increase a dataset to improve performance (16). Augmentations are therefore very commonly used on medical images, where datasets tend to be relatively small(15). Second, these transformations can help the network generalize better (15), as they alter features that are unimportant to the identification of COVID-19 in the lungs. This way the network can learn the important features and ignore the irrelevant ones. Crucially, the transformations must preserve the image labels - a coronavirus patient must still be identifiable as one. To ensure this, we consulted with radiologists when defining the transformations and their parameter ranges. The augmentations are performed randomly, with parameters chosen uniformly within the defined range as seen in Figure s3. Not all augmentations are applied each time, but rather each augmentation has a certain probability of being applied, represented by p below:

1. brighten, p=0.4
2. gamma contrast, p=0.3
3. CLAHE, p=0.4
4. rotate d ∈ [7,7] degrees p=0.4
5. shear d ∈ [7,7] degrees p=0.4
6. scale up to 0.2 on each axis p=0.4
7. flip from left to right, p=0.5
8. either sharpen or apply Gaussian blur
9. horizontal flip, p =0.5.

We decided to apply left to right flips, as COVID-19 is known to affect the lungs symmetrically. Thus, flipping will not change the characteristic manifestation of the disease. Moreover, some X-ray images may be taken from the back, and we do not always have clear labels as to the direction in which the X-ray was taken. Adding flips of the images can make the network robust to this.

**Figure s3:**
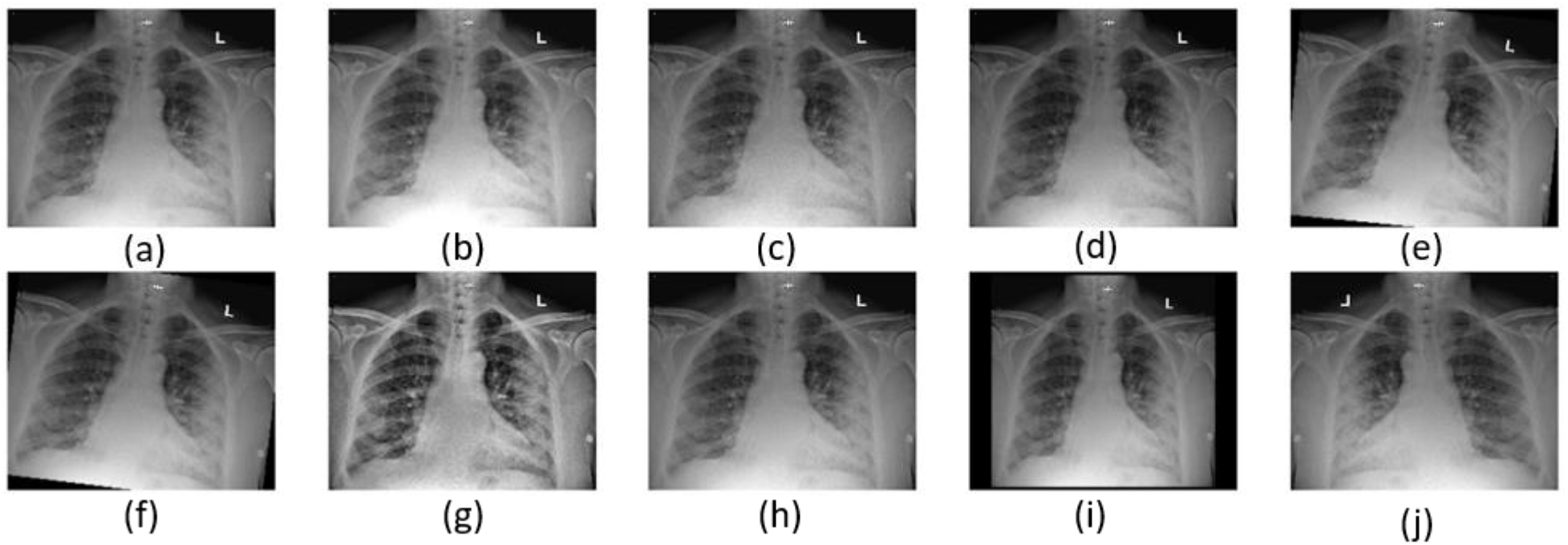
Image augmentation. In order to increase the number of images which can improve training performance, several different transformations are performed with a certain probability. The transformations showed: On top: (a) Original image, (b) Brighten, (c) Sharpen, (d) Gamma contrast, (e) Shear. On bottom: (f) Rotate 7 degrees, (g) CLAHE, (h) Gaussian blur, (i) Scale, (j) Horizontal flip.

### Segmentation

A novel aspect of our model architecture is adding an additional input channel to each image in the form of a probability vector, which indicates for each pixel the probability it belongs to the lung. These probabilities are obtained by applying a pre-trained U-net to segment the lung area from the image. Adding this mask as an additional channel to the X-ray image helps the network focus on the lung area while training. An example of segmentation can be seen in Figure s4.

**Figure s4:**
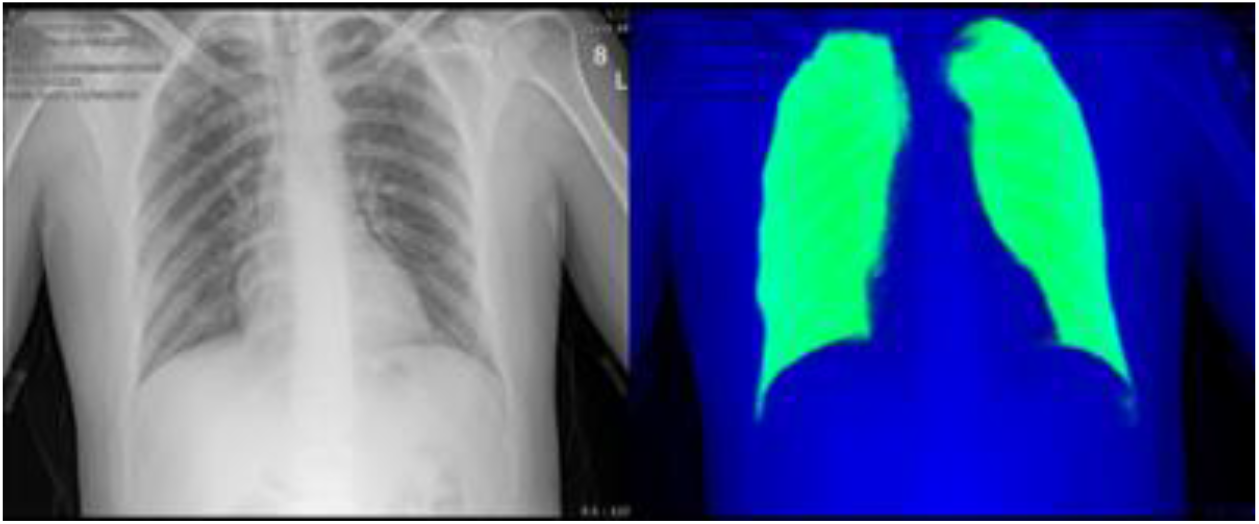
Lung segmentation using a U-net architecture.

### Network architecture

Deep learning-based automated diagnosis approaches have been gaining interest in recent years, mainly due to their ability to extract sophisticated features from images. This allows to describe an image in an alternative way from which we can derive computational conclusions. Based on that, our network architecture consists of two main parts - feature extraction, and decision head. The feature extractor is a neural network based on a Resnet50 architecture that gets an image as input (in our case - 2D image), performs mathematical operations on it and outputs a feature map, namely a matrix of numbers which describe the image. This matrix of features is converted to a vector (with the same values) and then goes into the decision head which is a simple neural network. In our case it consists of 3 fully connected layers. The output of the decision head is two numbers which describe the confidence of the algorithm about the classification results: COVID-positive or COVID-negative. In addition, the last layer (a vector) in the decision head is referred to as the “embedding” and is used as an input to the t-SNE and KNN algorithms described in the text.

**Figure s5:**
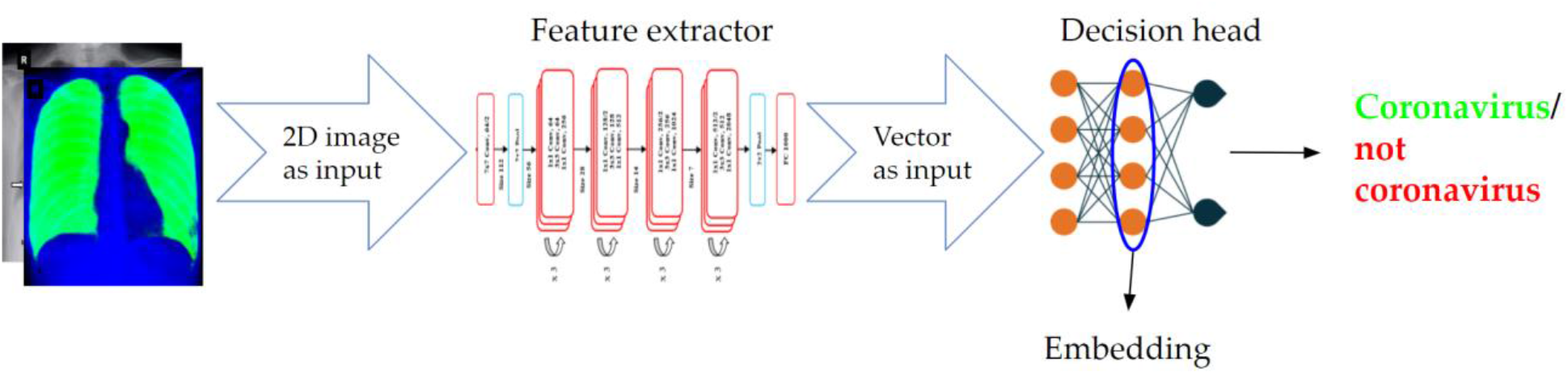
Pipeline of the neural network stage in inference time. Input images are passed through a sequence of convolutional layers that extract lower-dimensional vector representations for each image; these representations are optimized for the task at hand, in our case - separation in the vector space between images belonging to different label classes.

### Training details

Training was performed with the Adam optimizer with an initial learning rate of 1e-6 which was exponentially decreased as epochs progressed. We used cross-entropy as a loss function with an L2 regularization with regularization coefficient 1e-2. The best test accuracy scores were achieved after 32 epochs. The models were built and trained using Pytorch 1.6; all code will be made available upon publication.

### Bootstrap on the results

To analyze the stability and reliability of our results, we randomly split the data into train and test sets 10 times. In each split, the training set comprises 85% of the data, and each patient is used only for one of the sets.

For each split, we train the model on the train set, and then take 100 bootstrap samples from the corresponding test set, and compute the accuracy, specificity, sensitivity and AUROC for every bootstrap sample. Figure s6 shows that while there is some sensitivity to the way we split the data, most of the splits achieve similar results. The histograms show the distribution of scores obtained for each split. Split0 (in green) is the original split of the data that we worked with, and the one we reported all of our results on.

**Figure s6:**
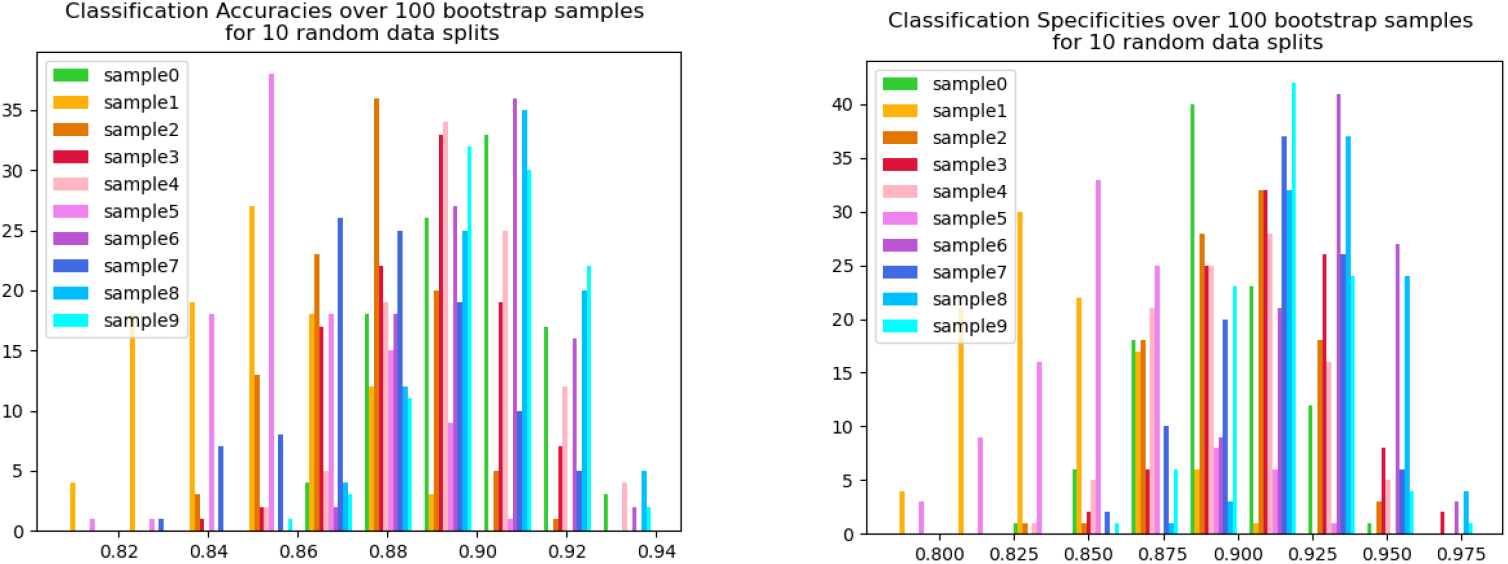
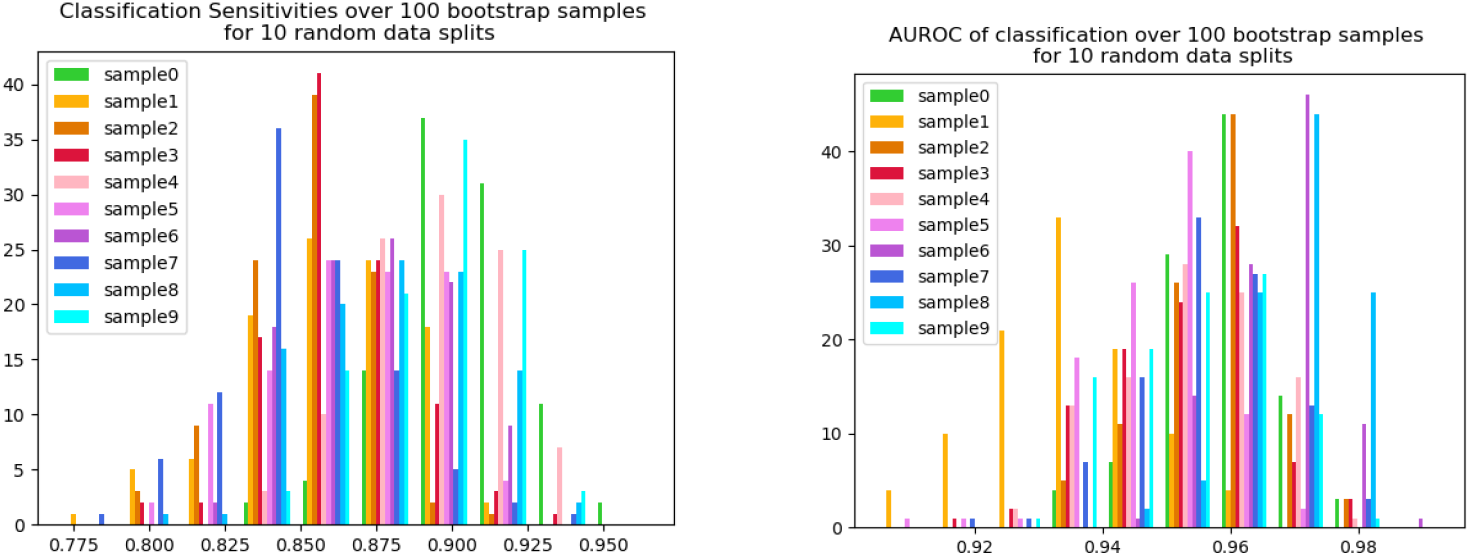
Distributions of results for100 bootstrap sampling for each one of 10 random splits of the data (a) Accuracy (b) Specifity (c) Sensitivity (d) Area under ROC curve.

## Notes

### Competing Interest Statement

The authors have declared no competing interest.

### Funding Statement

The work was funded by CoronaVirus Fund, Weizmann Insititue of Science.

### Author Declarations

This retrospective study was approved by the Institutional Review Board (IRB) and the Helsinki committee of the participating medical centers: 1) Department of Radiology, HaEmek Medical Center, Afula, Israel 2) Department of Otolaryngology, Head and Neck Surgery, Galilee Medical Center, Nahariya, Israel; 3) Cardiothoracic Imaging Unit, Shaare Zedek Medical Center, Jerusalem, Israel 4) Radiology department, Rabin Medical Center, Jabotinsky Rd 39, Petah Tikva; The study was approved in compliance with the public health regulations and provisions of the current harmonized international guidelines for good clinical practice (ICH-GCP) and in accordance with Helsinki principles. Informed consent was waived by the IRB of the above centers for the purpose of this study. Data extracted from medical records retrieved included only non-identifying information such as age, sex, vital signs, blood counts, chemistry, SARS-CoV-2 swab testing results, chemistry, and X-ray imaging files obtained as part of the diagnostic pipeline upon admission and on routine medical follow-up.

